# Reliability of serological tests for COVID-19: Comparison of three immunochromatography test kits for SARS-CoV-2 antibodies

**DOI:** 10.1101/2020.06.28.20140475

**Authors:** Hidetsugu Fujigaki, Masao Takemura, Michiko Osawa, Aki Sakurai, Kentaro Nakamoto, Koichi Setod, Takashi Fujita, Tadayoshi Hata, Hidehiko Akiyama, Yohei Doi, Kuniaki Saito

## Abstract

**Background:** Several immunochromatographic serological test kits have been developed to detect severe acute respiratory syndrome coronavirus 2 (SARS-CoV-2)-specific antibodies, but their relative performance and potential clinical utility is unclear.

**Methods:** Three commercially available serological test kits were evaluated using 99 serum samples collected from 29 patients diagnosed with coronavirus disease 2019 (COVID-19).

**Results:** The IgM antibody-positive rates of the three serological test kits for samples taken at the early stage of the disease (0–6 days after onset) were 19.0%, 23.8%, and 19.0%, respectively. The IgM antibody-positive rates over the entire period were 21.2%, 60.6%, and 15.2%, respectively. The IgG antibody-positive rates for samples taken after 13 days of onset were 100.0%, 97.6%, and 97.6%, respectively.

**Conclusion:** There were large differences among the results of the three test kits. Only few cases showed positive results for IgM in the early stage of disease and the IgM antibody-positive rates over the entire period were low, suggesting that the kits used in this study were unsuitable for diagnosis of COVID-19. The IgG antibody was positive in almost all samples after 13 days of onset, suggesting that it may be useful for determining infections in the recent past.

## 1. Introduction

Coronavirus disease 2019 (COVID-19) has been spreading globally. COVID-19 is caused by severe acute respiratory syndrome coronavirus 2 (SARS-CoV-2) infection, which was confirmed around December 2019 in Wuhan, Hubei, China [1, 2]. Currently, real-time reverse transcription-polymerase chain reaction (RT-PCR) is employed to detect SARS-CoV-2 in a nasopharyngeal swab or sputum for the diagnosis of COVID-19 [3]. However, RT-PCR gives false-negative results in cases of a low viral titer and inadequate sample collection. Studies in China using RT-PCR tests reported that only 60%–70% of COVID-19 patients were positive in the early stages of infection [3-6].

Detection of specific antibodies to a pathogen in the bloodstream is widely used to diagnose infectious diseases. An antibody test using a blood sample is relatively quick and straightforward, and because the risk of infection during the sample collection process is low, it is considered by some to be useful for the diagnosis of COVID-19 [7]. Generally, antigen-specific IgM antibodies increase in the early stage of the onset of a viral infection, which is then followed by an increased level of specific IgG antibodies. IgM antibodies are produced as the first antibody to fight the virus and are transiently raised. The subsequent production of IgG antibodies continues to rise for a long time and plays a vital role in immunity against the same virus. Therefore, the detection of specific IgM antibodies against SARS-CoV-2 may be used for diagnosis in the acute phase of COVID-19, whereas the detection of SARS-CoV-2-specific IgG antibodies may be used for determining a past infection or acquired immunity against SARS-CoV-2.

Immunochromatographic anti-SARS-CoV-2 antibody detection kits have been recently developed by multiple manufacturers [8]. However, differences in their properties and clinical usefulness are mostly unknown [9]. This study aimed to investigate the reliability of three different immunochromatographic anti-SARS-CoV-2 antibody detection kits using serum from COVID-19 patients.

## 2. Materials and Methods

### 2.1. Detection of anti-SARS-CoV-2 antibodies

We evaluated three test kits for the detection of anti-SARS-CoV-2 IgM/IgG antibody in serum: 2019-nCoV IgG/IgM Rapid Test Cassette (Hangzhou AllTest Biotech Co., Ltd., China), COVID-19 IgM/IgG Duo (SD BIOSENSOR, Korea), and 2019-nCoV IgG/IgM Detection Kit (Vazyme Biotech Co., Ltd., China). All kits were based on colloidal gold-labeled immunochromatography. All tests were performed following the manufacturer’s instructions for each kit, and the results were visually inspected within 15 min.

### 2.2. Study subjects

We utilized a series of residual serum samples left over after routine laboratory testing of 29 COVID-19 patients (mean age, 52.9 ± 21.9 years; 14 males and 15 females) who were admitted to Fujita Health University Hospital, Toyoake, Japan, from February 28 to April 15, 2020. All patients were confirmed as COVID-19 cases by RT-PCR assay of nasopharyngeal swab specimens at the time of or prior to admission. All serum samples, aliquoted and stored at −80°C, were thawed and evaluated at the same time for the analyses. This study was approved by the Ethics Committee for Clinical Research, Center for Research Promotion and Support, Fujita Health University (authorization number HM19-493).

## 3. Results

### 3.1. Positive rates of anti-SARS-CoV-2 IgM antibody

Table 1 shows the results of anti-SARS-CoV-2 IgM antibody in patients’ serum according to the number of days after disease onset. The IgM antibody-positive rate of the three kits in the early stage (0–6 days after onset) was 19.0% (Hangzhou AllTest), 23.8% (SD BIOSENSOR), and 19.0% (Vazyme Biotech), showing comparable results among the kits. On the other hand, the IgM antibody-positive rate 7 days after the onset showed considerable differences among the kits. After 13 days of onset, the IgM antibody-positive rates were 23.8%, 88.1%, and 14.3% for the Hangzhou AllTest, SD BIOSENSOR, and Vazyme kits, respectively. The SD BIOSENSOR kit showed a particularly high IgM antibody-positive rate (Table 1). The total IgM antibody-positive rates over the entire period were 21.2%, 60.6%, and 15.2% for the Hangzhou AllTest, SD BIOSENSOR, and Vazyme kits, respectively.

**Table 1.** Positive rate of anti-SARS-CoV-2 IgM antibody according to the different kits used.

| **IgM antibody** |  |  |  |  |  |  |  |
| --- | --- | --- | --- | --- | --- | --- | --- |
| **Days**<br>**after**<br>**onset** | No. of<br>samples | Hangzhou AllTest |  | SD BIOSENSOR |  | Vazyme |  |
|  |  | No. of<br>positive<br>samples | Positive<br>rate ( % ) | No. of<br>positive<br>samples | Positive<br>rate ( % ) | No. of<br>positive<br>samples | Positive<br>rate ( % ) |
| **0–6** | 21 | 4 | 19.0 | 5 | 23.8 | 4 | 19.0 |
| **7–8** | 10 | 1 | 10.0 | 2 | 20.0 | 0 | 0.0 |
| **9–12** | 26 | 6 | 23.1 | 16 | 61.5 | 5 | 19.2 |
| **After 13** | 42 | 10 | 23.8 | 37 | 88.1 | 6 | 14.3 |
| **Total** | 99 | 21 | 21.2 | 60 | 60.6 | 15 | 15.2 |

### 3.2. Positive rate of anti-SARS-CoV-2 IgG antibody

Table 2 shows the results of anti-SARS-CoV-2 IgG antibody in patients’ serum according to the number of days after onset. The IgG antibody-positive rates in the early stage (0–6 days after onset) were 33.3%, 33.3%, and 28.6% for the Hangzhou AllTest, SD BIOSENSOR, and Vazyme kits, respectively. Patients positive for IgG antibody were observed at the early stage of the onset.

**Table 2.** Positive rate of anti-SARS-CoV-2 IgG antibody according to the different kits used.

| **Days**<br>**after**<br>**onset** | No. of<br>samples | **IgG antibody** |  |  |  |  |  |
| --- | --- | --- | --- | --- | --- | --- | --- |
|  |  | Hangzhou AllTest |  | SD BIOSENSOR |  | Vazyme |  |
|  |  | No. of<br>positive<br>samples | Positive<br>rate ( % ) | No. of<br>positive<br>samples | Positive<br>rate ( % ) | No. of<br>positive<br>samples | Positive<br>rate ( % ) |
| **0–6** | 21 | 7 | 33.3 | 7 | 33.3 | 6 | 28.6 |
| **7–8** | 10 | 3 | 30.0 | 3 | 30.0 | 3 | 30.0 |
| **9–12** | 26 | 19 | 73.1 | 18 | 69.2 | 17 | 65.4 |
| **After 13** | 42 | 42 | 100.0 | 41 | 97.6 | 41 | 97.6 |
| **Total** | 99 | 71 | 71.7 | 69 | 69.7 | 67 | 67.7 |

The IgG antibody-positive rates after 13 days of the onset were 100%, 97.6%, and 97.6%, respectively, showing that the three kits gave positive results for almost all samples. Note that, since the IgM antibody-positive samples were also IgG antibody-positive, the positive rate of either or both IgM and IgG antibody was the same as the positive rate of the IgG antibody (Table 2).

### 3.3 Kinetics of anti-SARS-CoV-2 IgM and IgG antibody

Figures 1 and 2 show the kinetic results of anti-SARS-CoV-2 IgM and IgG antibody of the 29 patients in our cohort. Few cases showed positive test results for IgM early in the disease course. Tests for the IgG antibody often turned positive about 10 days after the onset, and most of the samples after 13 days of the onset became positive with all kits. However, there was a dissociation of the results in patient 11, where the AllTest kit gave positive results in all samples, whereas the SD BIOSENSOR and Vazyme kits gave negative results.

**Figure 1.**
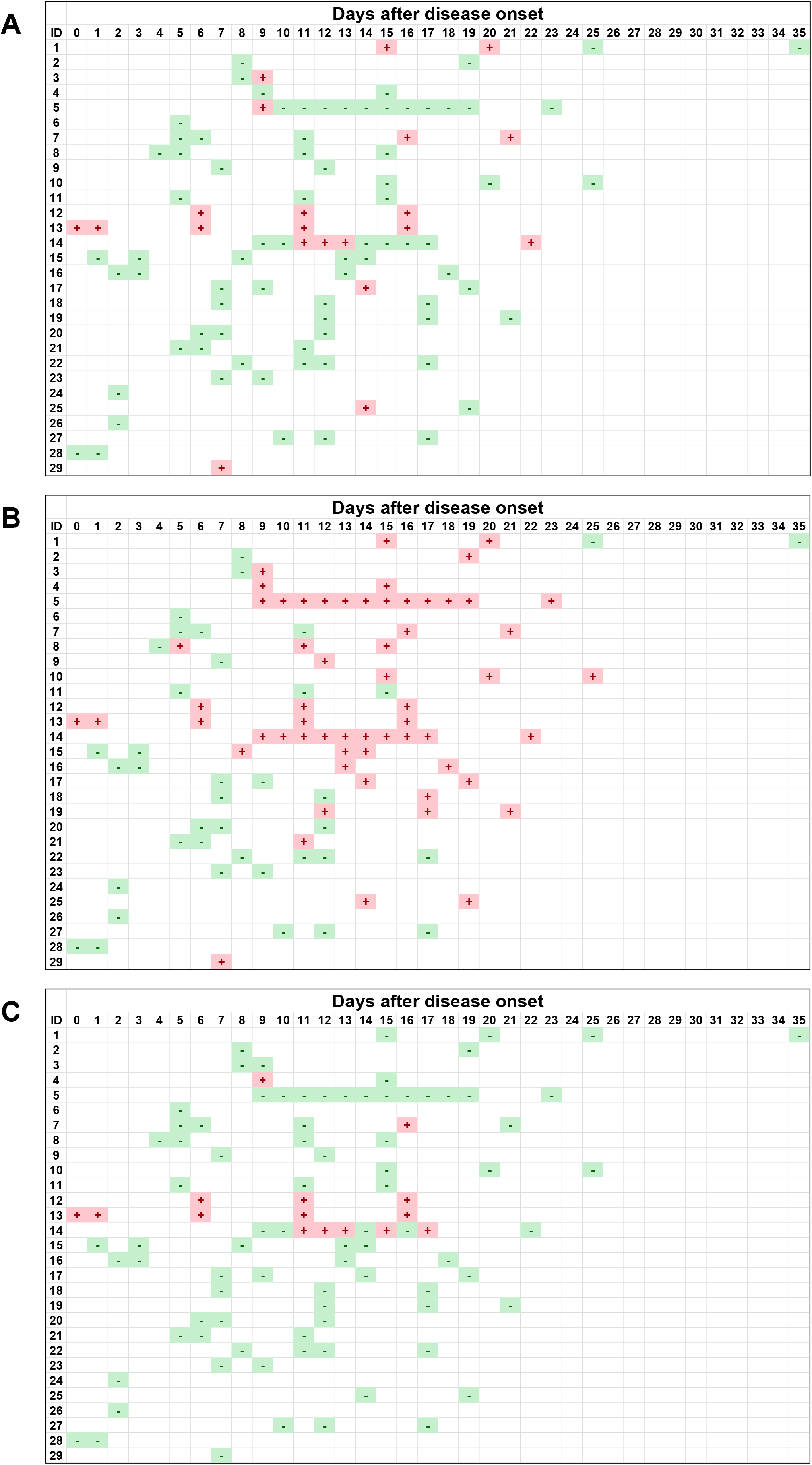
Kinetics of anti-SARS-CoV-2 IgM antibody of serum samples from 29 COVID-19 patients. The positive or negative result of the antibody in each serum sample is expressed as + or −, respectively. A, Hangzhou AllTest; B, SD BIOSENSOR; C, Vazyme.

**Figure 2.**
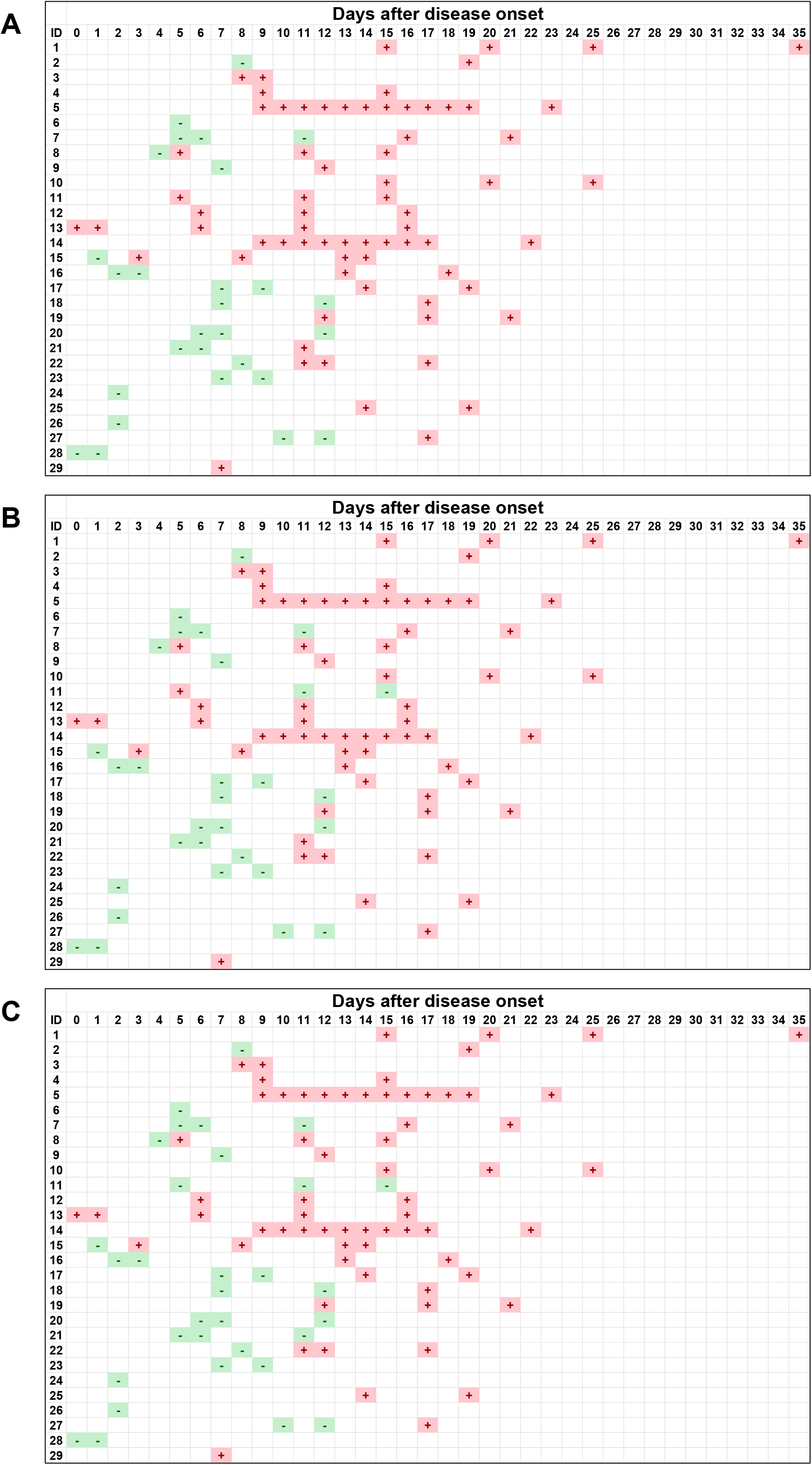
Kinetics of anti-SARS-CoV-2 IgG antibody of serum samples from 29 COVID-19 patients. The positive or negative result of the antibody in each serum sample is expressed as + or −, respectively. A, Hangzhou AllTest; B, SD BIOSENSOR; C, Vazyme.

## 4. Discussion

The serological detection of antibodies is widely used for the diagnosis of viral infections. Since immunochromatography methods to detect antibodies are quick to perform, easy to use, and do not require additional equipment, detection of anti-SARS-CoV-2 antibody using immunochromatography kits may be helpful for diagnosing COVID-19.

We compared three immunochromatography kits for detecting anti-SARS-CoV-2 antibodies using serum samples from COVID-19 patients. There was a considerable difference among the kit results, especially for IgM. It is thus clear that kit selection is crucial and should be based on the clinical purpose for the test. Additionally, since only a few samples were positive for the IgM antibody in the early stage of the disease by all kits, it appears that the kits used in this study are unsuitable for diagnosing the acute phase of COVID-19.

A comparison of the IgG antibody results between the three kits did not reveal any large differences. Additionally, almost all serum samples collected after 13 days of onset were positive. This suggests that the IgG antibody test using these kits can be helpful in diagnosing COVID-19 after a certain period from disease onset. However, the results of one patient were positive for one kit and negative for the other two kits, suggesting the presence of false-positive and false-negative results. Since research regarding the cause of false-positive and false-negative results in anti-SARS-CoV-2 antibody tests has yet to be performed, further investigation is necessary, such as determining the presence of cross-reactions with other coronaviruses. Furthermore, it is unclear how long anti-SARS-CoV-2 IgG antibody levels continue to rise in serum after infection, and further research is warranted.

Currently, epidemiological studies in several countries use immunochromatography kits to detect antibodies [10]. We suggest that when using immunochromatography kits for COVID-19 diagnosis, particular attention must be paid to whether these kits detect neutralizing antibodies against SARS-CoV-2. SARS-CoV-2 contains at least four structural proteins (spike [S] protein, envelope protein, membrane protein, and nucleocapsid [N] protein) [11, 12]. Since S protein appears to be involved in adhesion between virus and host cells during infection, it is considered that an antibody against S protein acts as a neutralizing antibody [13-15]. The detection of antibodies by immunochromatography kits may be useful for determinig a previous virus infection, but it is unclear whether the results could indicate people who have gained acquired immunity. Besides, quantitative assay of anti-SARS-CoV-2 antibodies, such as ELISA, would give a better understanding of humoral immune response to COVID-19 infection over time.

## Data Availability

The data that support the findings of this study are available from the corresponding author upon reasonable request.

## Acknowledgments

We would like to thank Nichirei Biosciences Inc. and Shionogi & Co., Ltd for providing the 2019-nCoV IgG/IgM Rapid Test Cassette and COVID-19 IgM/IgG Duo kits and the 2019-nCoV IgG/IgM Detection Kit, respectively.

